# A multi-modal phase plane method for constructing multivariate disease trajectories

**DOI:** 10.64898/2026.05.13.26353085

**Authors:** Timothy Cox, Rosita Shishegar, Pierrick Bourgeat, Marcela Cespedes, Vincent Doré, James D. Doecke, Jurgen Fripp, Christopher C. Rowe, Colin L. Masters, Victor L. Villemagne, Samantha Burnham

**Affiliations:** The Australian e-Health Research Centre, CSIRO, Melbourne, Australia; Department of Electrical and Computer Systems Eng, Monash University, Clayton, VIC, Australia; The Australian e-Health Research Centre, CSIRO, Herston, Australia; Department of Medicine, Austin Health, Heidelberg, VIC, Australia; Florey Institute of Neuroscience and Mental Health, The University of Melbourne, Parkville, VIC, Australia; Centre for Healthy Ageing, Health Future Institute, Murdoch University, Perth, WA, Australia; Department of Psychiatry, University of Pittsburgh, Pittsburgh, PA, USA

**Keywords:** Alzheimer’s disease, Longitudinal data, multivariate trajectories, biomarkers, disease course mapping

## Abstract

Understanding the sequential order and timing of different biomarkers in the progression of Alzheimer’s disease (AD) is paramount for understanding the pathophysiology of the disease, leading to better staging and improved prediction of clinical progression, providing crucial knowledge for the design and timing of effective clinical therapeutic trials. This study developed and evaluated a multi-modal phase plane (MMPP) method to construct long-term multivariate disease trajectory curves from short term longitudinal data for neuro-degenerative diseases like AD. The MMPP method is an extension to a previously presented four-step method for constructing single variable disease trajectories. A novel anchoring step which uses study participants’ multivariate data to infer the staging of the separate single variable progression trajectories allows multivariate disease trajectory curves to be generated. Further, the anchoring step provides disease staging at the individual level. A bootstrapping protocol was employed, providing confidence limits on the predictions. We demonstrate that the MMPP method is able to accurately reconstruct multivariate disease trajectory curves and individuals’ disease stage from simulated noisy short term longitudinal data. Specifically, the method successfully estimated the delay times between distinct progressing variables and reliably predicted individual baseline disease times (r^2^ = 0.981) for participants exhibiting significant early biomarker deviations.

## 1. Introduction

Alzheimer’s disease (AD) is the most prevalent form of dementia, accounting for approximately 70% of all cases globally *[1]*. Current consensus suggests that clinical interventions are most likely to succeed if administered during the pre-symptomatic phase, before significant neurodegeneration and clinical symptoms manifest *[2] [3] [4]*. Consequently, an in-depth understanding of the natural progression and pathophysiology of the disease is necessary to effectively design early-stage clinical trials *[2] [5] [6]*. This knowledge is essential for mapping the sequential order and timing of different biomarkers and for identifying the most appropriate individuals for trial recruitment or emerging disease-modifying treatments *[2] [7] [5] [3] [6] [4] [8]*.

Characterizing AD trajectories relies on sophisticated computational and statistical frameworks designed to bridge short-term patient observations with the multi-decade progression of the disease. These approaches leverage large-scale longitudinal datasets to synthesize long-term timelines from localized, cross-sectional biomarker measurements. A critical challenge in this domain is the temporal alignment problem: the necessity of overcoming inter-subject variability to convert isolated data points into a continuous, universal “disease age” or “biological clock” *[5] [3] [4]*.

Neocortical Amyloid-beta (Aβ) plaque, measurable via positron emission tomography (PET), is among the earliest and most studied biomarkers of the preclinical phase of AD. In previous work, we developed the phase plane method, a four-stage statistical framework designed to elucidate the natural history of Aβ accumulation using PET imaging data from the Australian Imaging Biomarkers & Lifestyle (AIBL) study and the Alzheimer’s Disease Neuroimaging Initiative (ADNI) [2]. This approach demonstrated that Aβ deposition is a prolonged process spanning approximately 30 years, highlighting a substantial preclinical window that has informed secondary prevention trials [2]. Furthermore, it revealed that APOE ε4 allele carriers begin accumulating approximately 15 years earlier than non-carriers [2].

Despite the central role of Aβ, it represents only one component of an evolving biomarker landscape. Additional markers, including Aβ42 in cerebrospinal fluid (CSF) and plasma, tau pathology, astrocytic function (glial fibrillary acidic protein, GFAP), neurodegeneration (neurofilament light (NfL and MRI-derived measures), and measures of cognitive and functional decline, collectively provide a more comprehensive characterization of progression. Understanding the temporal dynamics and interactions among these multifaceted markers is critical for clinical applications, such as determining how many years cognitive deterioration is preceded by specific pathological events [9] [5]. To capture the proposed “S-shaped” (sigmoidal) dynamics, researchers employ various non-linear frameworks [9] [5] [6]. Foundational studies initially utilized simple linear regression slopes mapped against baseline values to integrate average rates of change over time [2]. More recently, Accelerated Failure Time (AFT) models allow for the estimation of individual “temporal shifts” to identify lags between markers, such as the rise of GFAP relative to phosphorylated tau (pTau) [6]. Furthermore, mixed-effects models combined with quantile regression and natural or restricted cubic splines are used to define the timing between milestone events by robustly capturing subject-specific non-linearities [9] [5].

While existing disease progression models have made important contributions, many remain limited to single variable formulations and rely heavily on strict biological thresholds. Recent advancements have introduced analytical “clock models” to align individual timelines to estimated symptom onset using biomarker positivity *[3] [4]*. Methods such as Sampled Iterative Local Approximation (SILA) act as non-parametric algorithms that iteratively sample and “stitch” together discrete longitudinal snapshots to infer a population trajectory without assuming an underlying mathematical shape *[10] [4] [8]*. Conversely, models such as Temporal Integration of Rate Accumulation (TIRA) *[4]* and ordinary differential equation–Gaussian process (ODE–GP) *[10]* return to phase-plane principles, mathematically integrating the inverse relationship between robustly modeled rates of change and current physiological values to derive a disease timeline. Such sophisticated non-linear methods are inherently “data-hungry,” and their successful implementation relies on the massive longitudinal datasets now available through global consortia or harmonised datasets *[11] [12] [13] [14]* to accurately model individual-level variations in disease evolution. Foremost, while TIRA, (ODE–GP) and SILA leverage powerful statistical machinery, they are fundamentally designed to evaluate individual biomarkers in isolation *[10] [4]*.

In this work, we extend our original phase plane method to a multi-modal setting, introducing the multi-modal phase plane method (MMPP). Building upon foundational phase-plane integration *[2]*, this framework enables the simultaneous modelling of multiple biomarkers by introducing a novel global optimization step that dynamically shifts and anchors multiple distinct biomarker curves for each individual onto a single, universal “disease time” axis. By facilitating the estimation of an individual’s holistic position within the multi-modal disease course, MMPP supports personalized staging, prognosis, and the identification of candidates for targeted clinical trials and therapeutic interventions.

## 2. Methods

### 2.1 Method Overview

Here we propose the MMPP method, a 5-step procedure, Figure 1, to obtain overall disease progression for a multi-modal set of biomarkers. Single trajectory disease curves are constructed using the four-step method [2] [7], previously proposed by our group. Here, mean trajectory curves *μ*_*Q*_(*τ*) for each variable of interest, *Q* are obtained. This is achieved by: 1) obtaining a series of points in the *phase plane* consisting of rate of change estimates and mean values for each of the variables for each participant, 2) estimate the rates of change of the mean trajectories 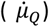 as a function of *μ*_*Q*_, 3) obtain disease time variables (*τ*_*Q*_, measured in years) as a function of *μ*_*Q*_, 4) invert this relationship to get the single variable mean trajectory curves *μ*_*Q*_(*τ*_*Q*_) as a function of disease time. 5) The overall disease time *τ* for the multi-modal variables is then determined using independent time shift constants or *anchoring times* for each multimodal variable. To achieve this the discrepancy between all individual’s multivariate longitudinal data and the constructed disease trajectory curves are minimised. The minimisation procedure not only provides the anchoring time for the different curves and staging of the multivariate curves, but it also gives an estimate for where each participant’s multivariate data lies on the trajectory.

**Figure 1:**
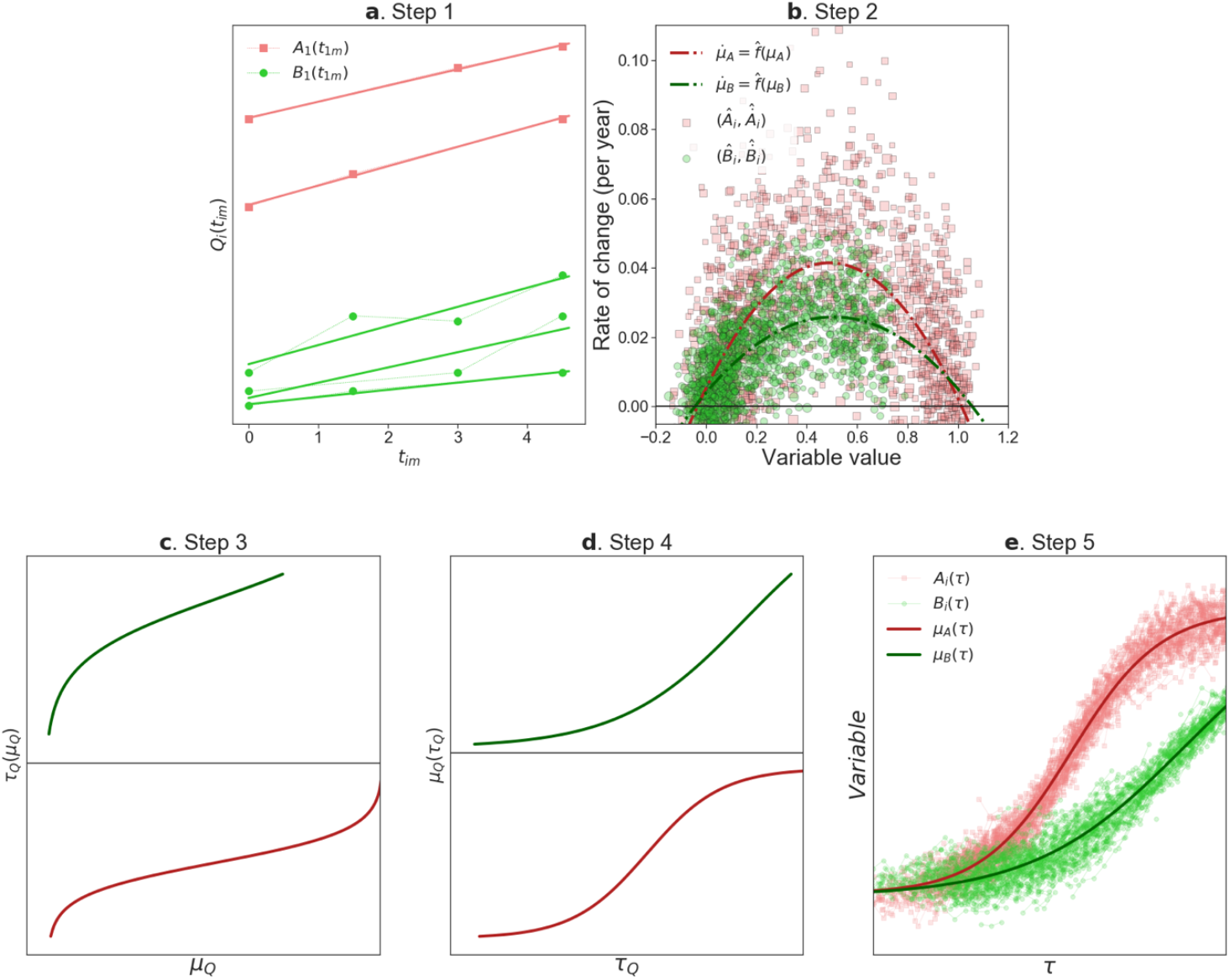
The five step process for two biomarkers shown in red and green: **a** Step 1, linear rates of change of the individual variables are estimated from the longitudinal data, this example is shown for five participants. **b** Step 2, the functional relationship between the rates of change of the variables are determined by fitting to the rate of change data. **c** Step 3, a disease time is determined for each variable as a function of the variable by integrating the fit curves. **d** Step 4, the relationship is inverted to give each variable as a function of its disease time. **e** Step 5, the multivariate longitudinal data is then used to determine the relationships between the different disease times and each individual’s disease time.

Given longitudinal data on a set of quantities (biomarkers or other quantities) 𝒬 measured for *i* = 1,2, …, *N* individuals. Let *t*_*im*_ be the time since baseline of the *m*’th follow up observation of the quantities for the *i*’th individual measured after their first observation (at *t*_*i*0_ = 0). We denote the observed value for the quantity Q for the individual *i* at the collection time *t*_*im*_, by *Q*_*i*_(*t*_*im*_).

The key assumption is that the trajectories of the quantities for any individual are well approximated by a multivariate mean trajectory, (which consists of a set of single variable trajectories {*μ*_*Q*_(*τ*) for *Q* ∈ 𝒬}) with a time shift, equation (1),

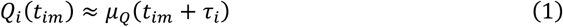

where the *τ*_*i*_ represents an anchor time (or time shift), which specifies how far the disease has progressed for the individual *i* at the time of their first observation (and *t*_*im*_ + *τ*_*i*_ is the individual’s disease time at their *m*’th data collection). To avoid confusion over the different “time variables” used in this manuscript, the time variables are summarised in Table 1. The goal of this study is to construct estimates of the multivariate mean trajectory and individual anchoring times.

**Table 1:** Explanation of the different time parameters estimated via the multivariate approach (all time variables are measured in years).

| Symbol | Description | Text section |
| --- | --- | --- |
| $\tau$ | *Disease time*: a measure of disease progression. Defined as the dependent variable of the multivariate mean trajectories $\mu_Q(\tau)$. $\tau$ is defined such that when $\tau = \tau_{Q0}$ $\mu_Q$ is at the cut-off value for $Q$ (equation 3). | Equation (1),<br>Section 2.1 |
| $t_{im}$ | Time of $m$'th longitudinal data point for participant $i$. | Section 2.1 |
| $\tau_i$ | *Anchor time* for individual $i$, the disease time of the participant $i$'s baseline measurement. Provides an individualized measure of disease progression. | Section 2.1 |
| $\tau_Q$ | *Single variable disease time* for the variable $Q$, parametrises the single variable disease trajectories $\mu_Q^*(\tau_Q)$, obtained via the four-step method. | Section 2.1 |
| $\tau_{Q0}$ | *Anchor time* for variable $Q$, relates the single variable disease time variables from four step method to the disease time $\tau = \tau_Q + \tau_{Q0}$. | Equation (3),<br>Section 2.1 |
| $\theta_{im}$ | *Simulated disease time* for simulated individual $i$ at time $t_{im}$ | Equation (6),<br>Section 2.2 |
| $\tau_{Q0b}$ | The value of $\tau_{Q0}$ computed in bootstrapped replica $b$ | Section 2.6 |
| $\tau_{ib}$ | The anchor time for individual $i$, in the bootstrapped replica $b$. | Section 2.6 |
| $T_b$ | The time shift for bootstrapped replica $b$ | Section 2.6 |
| $\tau_b$ | The disease time for bootstrapped replica $b$ | Section 2.6 |
| $\tau_{Q_{1stb}b0}$ | The disease time at the earliest cut-off point in bootstrapped replica b | Section 2.6.1 |

To estimate the multivariate mean trajectory, first we estimate single variable mean trajectory 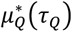 for each variable using the four-step approach developed by, Budgeon *et. al*. [7] and Villemagne *et. al*. [2], which consists of the following steps:

#### Step 1

Estimated phase plane points 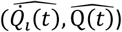, consisting of an estimate for the rate of change of the variable and the mean value of the variable are calculated from each individual’s longitudinal data (we use the notation 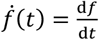 for time derivatives). This step is, described in section 2.3.

#### Step 2

An estimate of a function

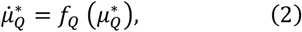

which provides the rate of change of the variable as a function of the mean value of the variable is obtained by fitting a function to the phase plane points, as discussed in 2.4.

#### Step 3-4

Solve the differential equation (2) to obtain 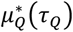, which includes an arbitrary integration constant. This is discussed in section 2.4.

The four-step method leaves us with a set of mean single variable trajectory curves {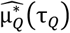 for *Q* ∈ 𝒬}, which are related by a set of anchoring times {*τ*_*Q*0_ for *Q* ∈ 𝒬} to the best estimates of the multi-variate trajectory curves {μ_*Q*_for *Q* ∈ 𝒬},

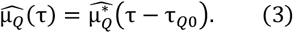

To derive a zero-point disease time, pre-defined single variable cut-off values *Q*_*c*ut–off_ are used for each biomarker. Mathematically we set

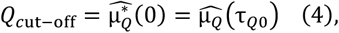

such that we can write the specific solution to the differential equation (3)

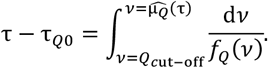

In order to estimate anchoring times, we add a novel step, which allows the multivariate trajectories to be constructed from the single variable trajectories for the first time:

#### Step 5

Estimates for a set of anchoring times {*τ*_*i*_ for *i* ∈ {1,2, …, *N*}} (one for each individual) and {*τ*_*Q*0_ for *Q* ∈ 𝒬} (one for each biomarker) are produced, by minimising a weighted error between the μ_*Q*_ curves and the longitudinal multivariate data, this is introduced in detail in Section 2.5.

### 2.2 Simulated Data

Simulated longitudinal data for two variables, *A* and *B* (so that 𝒬 = {*A, B*}) is utilised to demonstrate and test the method. The five-parameter logistic functions described in Gottscahalk and Dunn [15] are employed to generate the simulated data where values for individual *i* at times *t*_*im*_ are,

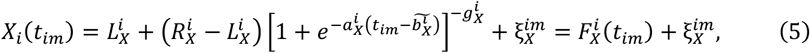

Here *X* ∈ {*A, B*} specifies the variable, 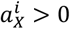 specifies how rapidly the variable increases, while

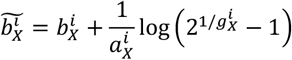

is the time at which the variable is halfway between the two asymptotes of the curve; 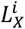 defines the variable’s lower asymptote; 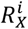 defines the variable’s upper (and right) asymptote; 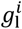 determines the asymmetry of the curve (when 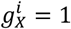 the curve is symmetric around its mid-point); 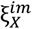 represents a random addition to the *m*’th observation of the variable; and 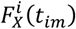 is the part of the function that doesn’t depend on the random “noise”. This gives simulated trajectories (Figure 2), with an asymmetric sigmoidal shape similar to those in Budgeon *et. al*. [7], but with the addition of point noise.

**Figure 2:**
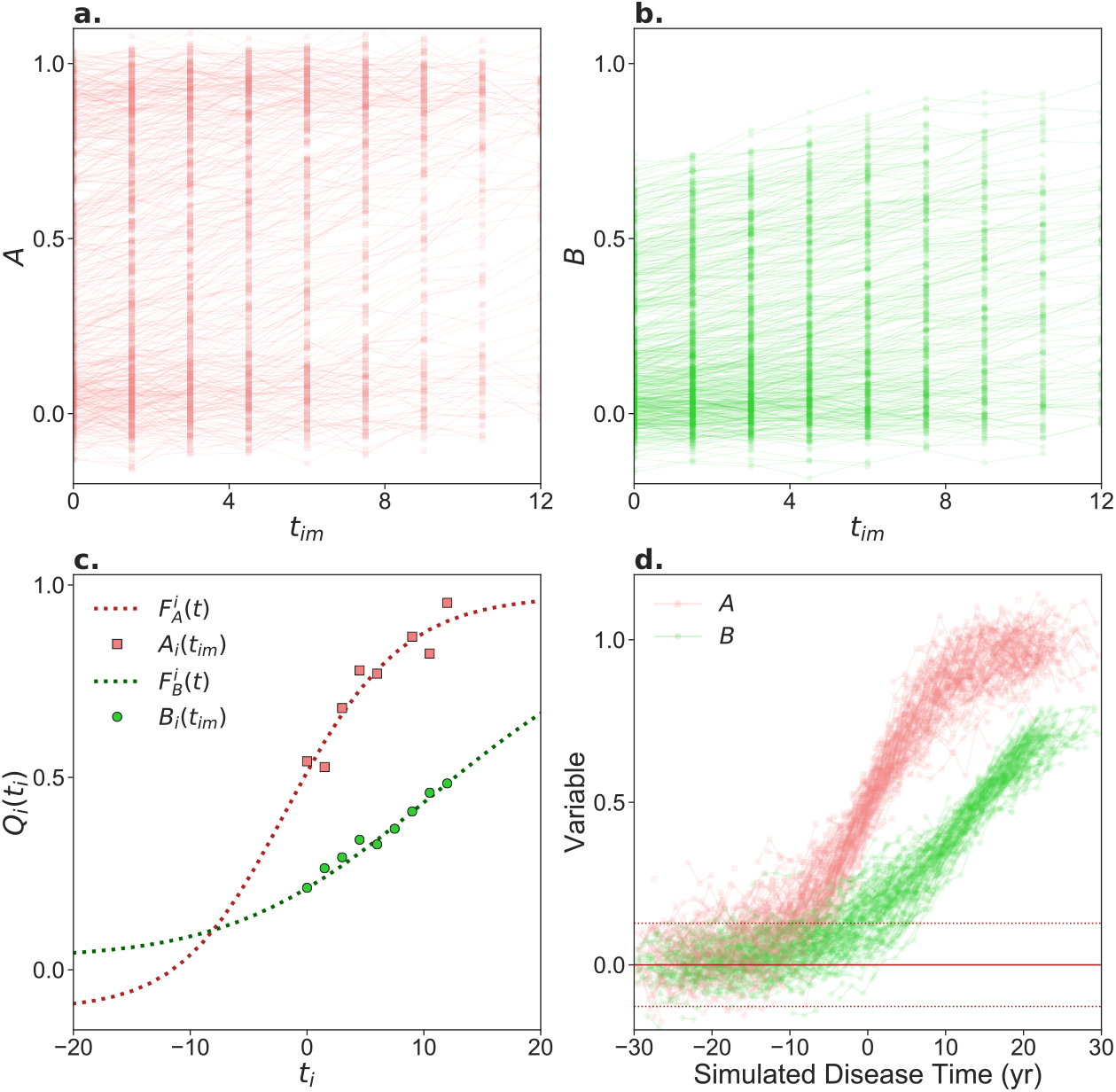
a & b The full simulated longitudinal data set for the variable *A* (& *B*). **c** The simulated longitudinal data for a single simulated individual. The 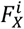 curves are shown with lines and the simulated data points are shown with points. The curve parameters for this individual are as follows, *A:* Curve parameters: 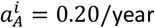, 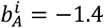 year, 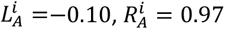, and 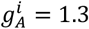, and *B:* Curve parameters: 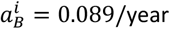, 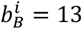 year, 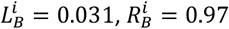, and 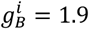. **d** The full simulated longitudinal data set for the variables *A* and *B* plotted as a function of the simulated disease time variable, the simulated pre-disease mean of the *A* variable is shown with a solid line, and the two standard deviations spread of the pre-disease population are marked with dotted lines.

#### Simulated dataset details

We generate the example data set from equation (5) using the following parameters for each of *N* = 1200 simulated individuals with the following independent distributions,

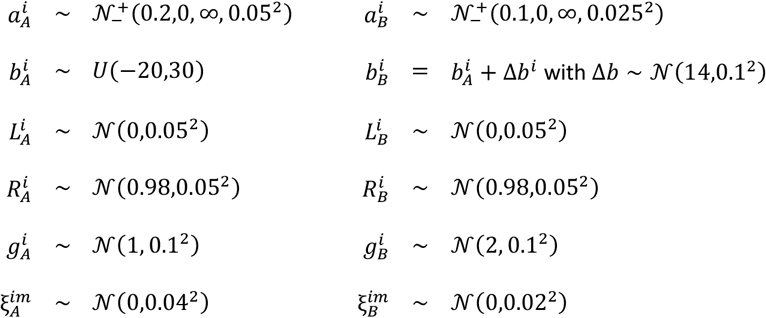

where 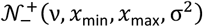 is the truncated normal distribution, obtained by truncating the normal distribution 𝒩 (ν, σ^2^), to the interval [*x*_min_, *x*_max_]. These parameters have been selected to simulate a realistic disease progression of two variables, where on average the *B* curves increase slower than A and have a lag of ∼*14 years*. Each individual’s simulated longitudinal data consists of *n*_*i*_ data points at times 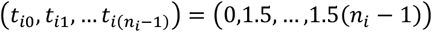, where the number of points for each individual are randomly generated using a discreate uniform distribution 𝒰, *n*_*i*_ ∼ 𝒰{1,9}, and the variables are calculated using equation (2). To simulate missing at random values common in real datasets, we remove 10% of the data points for each variable at random. An example of a simulated individual’s data and the full simulated data set is shown in Figure 2.

#### Simulated disease time

To test how well the method places individual study participants on the multivariate trajectories, a measure of how far the simulated longitudinal study participants are along the disease trajectory curve is needed. To this end, we define the simulated disease time *θ*_*im*_ for a simulated individual *i* at time *t*_*im*_ by

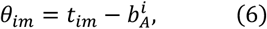

this definition ensures that a simulated participant’s *A* variable will be half-way between its final and initial values at *θ*_*im*_. The dependance of the variables *A* and *B* for the entire simulated data set are shown in Figure 2.

### 2.3 Step 1: Calculating phase plane points from single variable longitudinal data

Step 1 uses a set of points for each participant to calculate both the estimate of the rate of change of each individual’s variable 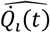, and an estimate of the concurrent value of that variable 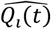; these define points in the *phase plane* for each variable.

When estimating phase plane points Budgeon et. al. [7] used a method based on an ordinary least squares linear fit to the full data for each individual to compute the estimate 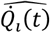, and the mean biomarker data for each individual to estimate 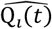. Note the accuracy of these estimates depends on the number of data points for each individual and the time these data points spanned.

### 2.4 Steps 2-4: Fitting the phase plane data & solving for the single variable trajectory curves

After the phase plane data has been calculated, we fit a function *f*_*Q*_(μ_*Q*_) for each biomarkers’ phase plane data, so that each μ_*Q*_ satisfies the differential equation (2). We fit parametrised functions *f*_*Q*_(μ_*Q*_, **β**) by adjusting the parameters, **β** to minimise the Mean Squared Error, (MSE),

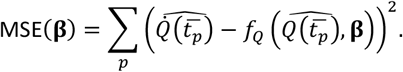

We use least squares polynomial fits for fitting the phase plane data in the remainder of this study. The choice of the polynomial order is discussed in [7], in line with this we utilise the lowest order polynomial which accurately fits the data. The polynomial fit to our simulated data is shown in Figure 1a, where we use a second order polynomial.

The third and fourth steps solve the differential equation (2) for each of the single variable trajectory curves. This is done first by integrating the differential equation to get the disease time as a function of the single variable trajectory (step 3), then inverting the relationship gives the single variable trajectory 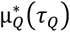 as a function of a single varable disease time *τ*_*Q*_ (step 4). At the end of step 4 we have a set of single trajectory curves with their own disease time *τ*_*Q*_ as shown (for our simulated data set) in figure 1 d.

Each integration in step 3 introduces an unknown integration constant, which can be interpreted as a time shift *τ*_*Q*0_ such that the multivariate trajectories can be written as follows,

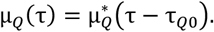

The anchor times {*τ*_*Q*0_ for *Q* ∈ 𝒬} are determined in the next step. Numerical values of 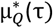 are computed by standard methods as described in section A of the supplementary material.

### 2.5 Step 4: Calculating the anchoring times

In order to derive a maximum likelihood estimate for the undetermined anchoring times {*τ*_*Q*0_} we assume that each participant’s biomarker values are normally distributed around some point on the multivariate trajectory, that is for the participant *i*

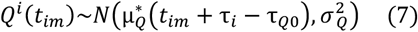

where *τ*_*i*_ is the *anchor time* for the for the individual *i* and σ_*Q*_is the standard deviation for the biomarker *Q*. The anchor time for an individual can be interpreted as the disease time at baseline at baseline for that individual. The anchor times for all biomarkers {*τ*_*Q*0_ for *Q* ∈ 𝒬} and individuals {*τ*_*i*_ for *i* ∈ {1,2. ., *N*}} as well as the standard deviations {σ_*Q*_ for *Q* ∈ 𝒬} can be determined by minimizing the logarithm of the total likelihood ℒ ({*τ*_*i*_}, |*τ*_*Q*0_}, |σ_*Q*_}),

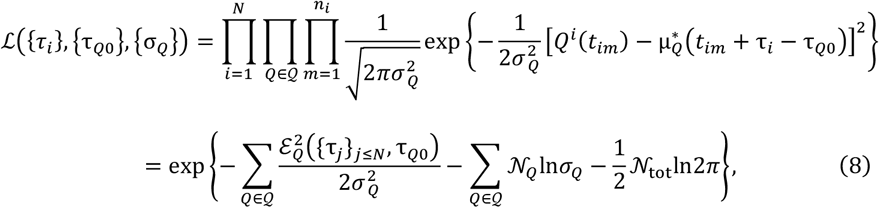

where 𝒩 _*Q*_is the total number of values for the biomarker *Q*; 𝒩 _tot_ = ∑_*Q*_ 𝒩 _*Q*_ is the total number of datapoints, and 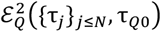 is the summed squared error for Q,

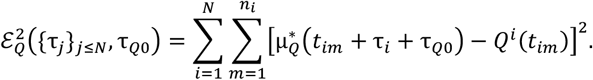

The likelihood function (8) will not have a unique minimum, as adding an arbitrary constant to all the anchoring times leaves the likelihood unchanged. To remedy this, we fix one of the *τ*_*Q*0_ = 0. The standard deviations σ_*Q*_ can be eliminated by solving 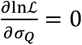, which gives

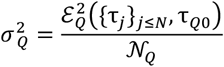

Substituting the 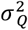 back into the likelihood and taking logarithms gives,

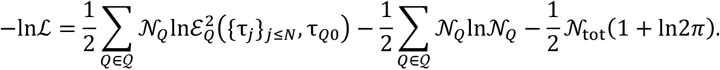

The final two terms are constants which leaves use with the loss function −*ℓ* to be minimised,

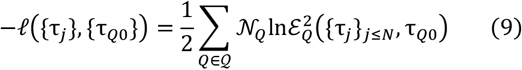

Minimising −*ℓ* to provides maximum likelihood estimates for the anchoring times.

#### Minimisation algorithm

We minimize the negative log pseudo likelihood (9) using the Broyden–Fletcher–Goldfarb–Shanno (BFGS) quasi-Newton algorithm, as implemented in SciPy. This algorithm requires an initial guess and that the problem is regularized such that participants who have longitudinal data on the flat parts of the curve have a definite placement. Both the regularization procedure and the construction of the initial guess are described in sections B and C of the supplementary material respectively.

### 2.6 Bootstrapping for confidence estimation

A bootstrap methodology was used to estimate confidence intervals for the various quantities calculated with the five-step method (disease trajectories, disease times for individuals etc.). The bootstrapping method is a modified version of that described by Budgeon et. al. [7], with the modifications accounting for the extra complexity of multivariate disease information and individual anchor times. We produce *N*_*B*_ bootstrap replicas of our longitudinal data set ℒ by sampling (without replacement) *N*_Ind_ individuals in the longitudinal data set. This produces *N*_*B*_ longitudinal sets 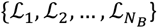. Performing the five step method as described in sections 2.3-2.5 on each of data sets produces a set of multivariate disease curves 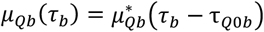, anchor times *τ*_*ib*_, and different *τ*_*b*_ for each bootstrap replicate. For each replicate the anchoring times are set up-to an arbitrary constant time shift. To make this explicit we set *τ*_*b*_ = *τ* + *T*_*b*_ where *τ* is a disease time which is comparable between different bootstraps. In general, the user will have a choice of *T*_*b*_, depending on the biological question being asked. For example, if the user was interested in the population level dependence of participants who started at a fixed value of the marker *A* = *A*_milestone_, *T*_*b*_ values can be fixed such that every curve went through *A* = *A*_milestone_ at disease time zero.

In the results section, since we are interested in the participants disease trajectory and the general shape of the disease curves, we fix the *T*_*b*_ variables by minimising the sum of the variances of each individual’s time shift. We do this minimisation with SciPy’s least_squares function and an initial guess of all *T*_*b*_ = 0.

In general the user will be interested in using the curves to map the average trajectory of participants AD progression (this is encapsulated in the assumption of equation (1)) at the typical age amongst the progressing participants. This suggests obtaining *T*_*b*_ values by setting them to the average age that the first of the markers reaches abnormal values, this is described in more details in the next section and leads to the proposed disease age.

It is worth noting that quantities depending on disease time difference do not require the fixing of the *T*_*b*_ quantities to obtain bootstrapped confidence estimates of some results. An example of such a quantity would be the difference in time between two variables reaching statistically significant values compared to a pre-disease population.

#### 2.6.1 Computing disease age

For each bootstrap, we determine which of the cut-off values occurs first on the disease trajectory, that is the biomarker *Q*_1stb_ for which in bootstrap b, 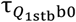 (the anchor time & time at cut-off value for *Q*_1stb_) is smallest. The baseline time since this first threshold for the participant *i* is then (compare equations (3) and (7))

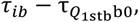

where *τ*_*ib*_ is the anchor time for the participant *i* in that bootstrap. If the baseline age for this participant is (age)_*i*_ then an estimate for age at that first cut-off (A1CO) is

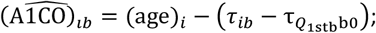

this is illustrated for a single example participant in Figure 5a & c. In general, we would only expect 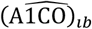 estimates to be accurate for participants who are passed the first cut-off value to match the cut off time with the median 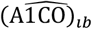 that is,

**Figure 4:**
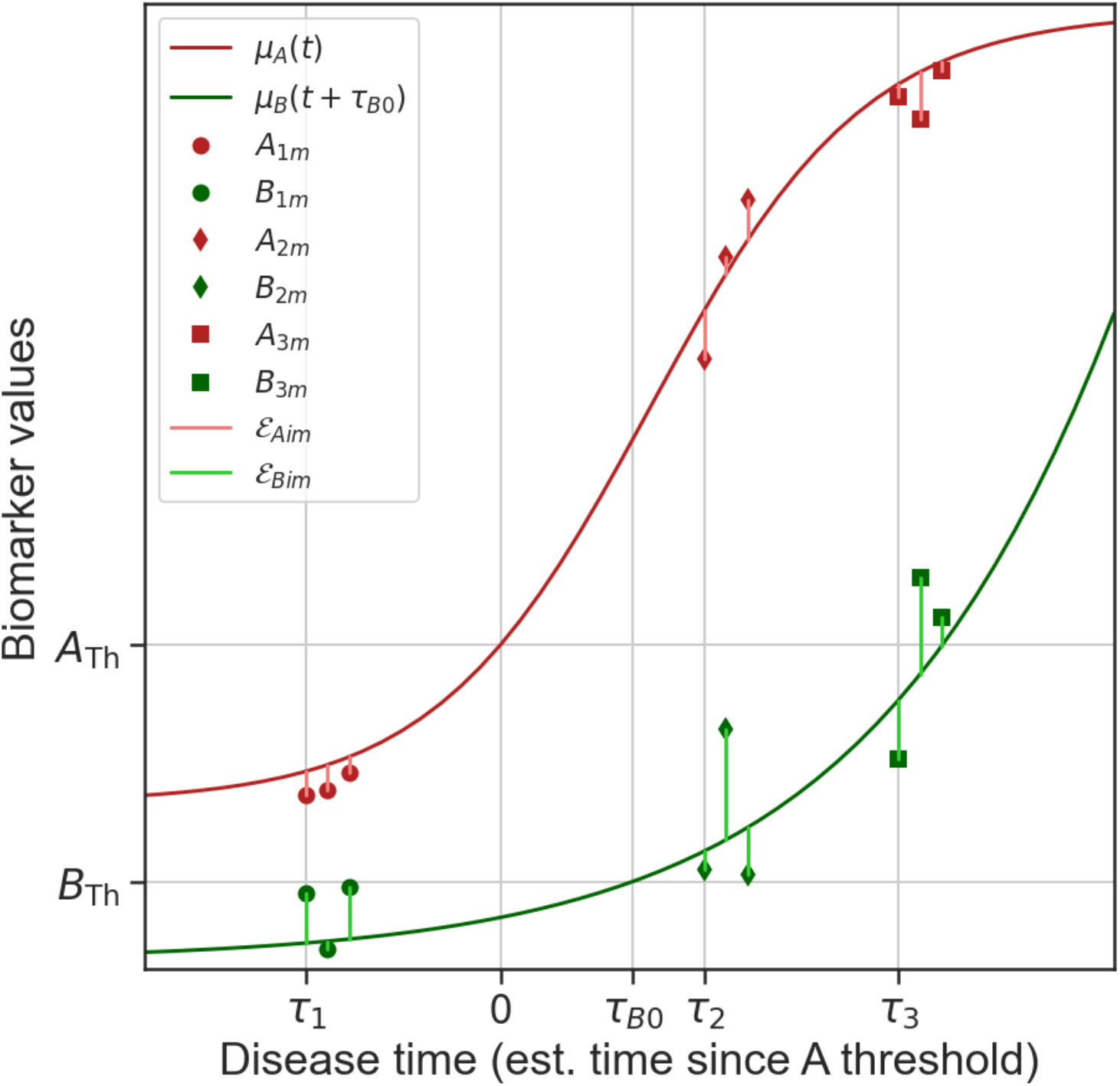
Step 5 Obtaining the anchoring times by minimizing the error. Both trajectory curves are shown (dash dotted lines) as well as the longitudinal data points (for three simulated participants) obtained after the anchoring points have been fit by minimizing the weighted error as discussed in the text. The error for each point is illustrated by the vertical lines, while the dotted lines connect single variable longitudinal trajectories for each individual.

**Figure 5:**
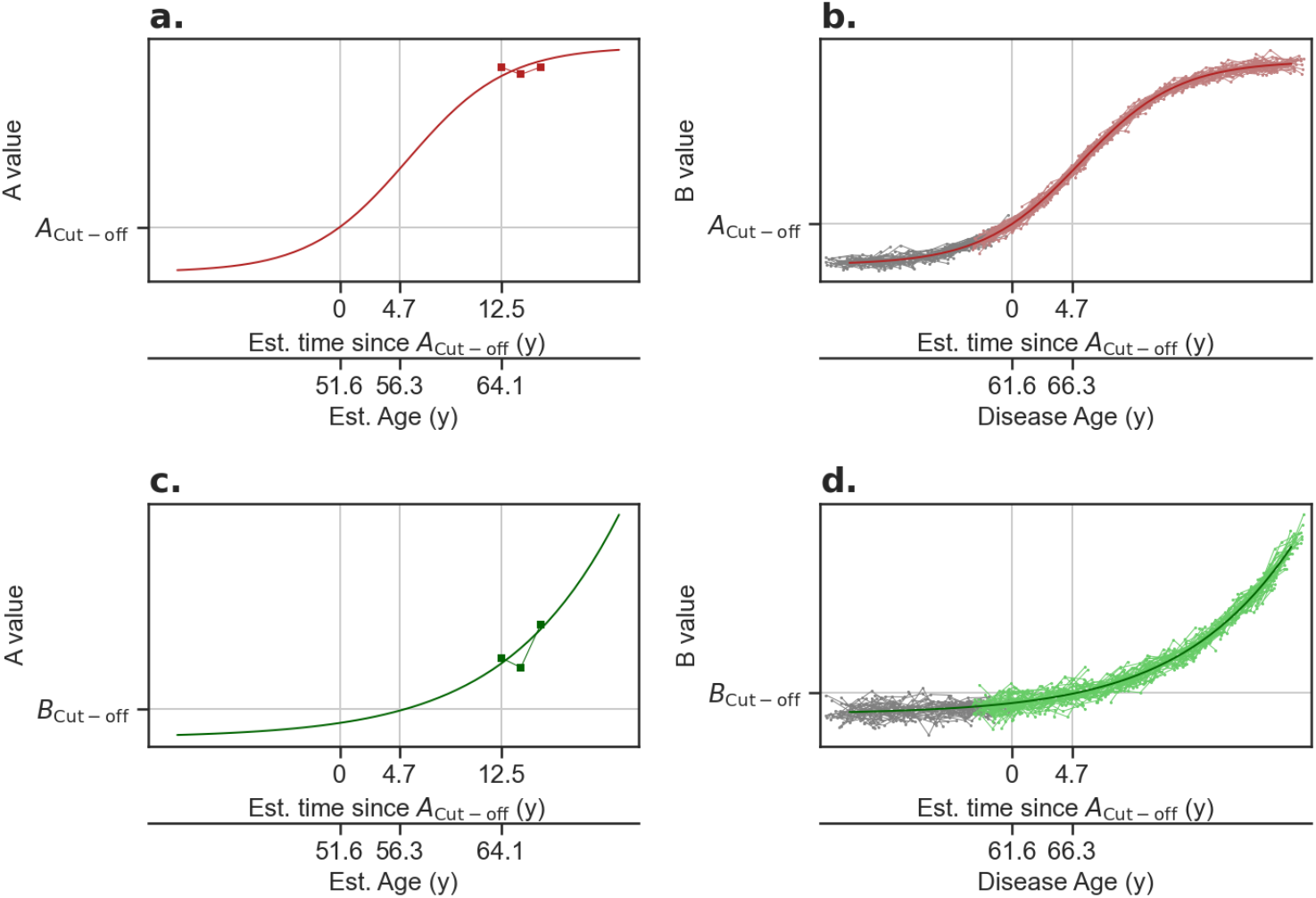
disease age computation for a two biomarker example, a & c the disease curves for the biomarkers A & B and placement for a participant *i* with (*age*)_*i*_ = 64.1 y and *τ*_*i*_ − *τ*_*A*0_ = 12.5 *y* (the biomarker A reaches its cut-off first in this example), for this participant the estimated age at first cut-off is (*A*1*CO*)_*i*_ = 51.6 y. Solid line is the disease curve; the connected points show one participants data, and the horizontal axes show both estimated time since cut-off for A (disease time), and estimated age for that participant. b & d illustrate the computation of the estimated age at first cut-off and the disease age. The connected points are participants data anchored on the curves participants with coloured data have at least one point with disease time (estimated time since A cut-off) greater than zero one the median of the estimated age at A cut-off among those participants is taken on gets the median 61.6 y, so that the disease age scale has a value of 61.6 y when A is at its cut-off. The dark curve shows the disease time.

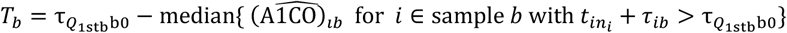

(the median is taken over all participant with a final disease time 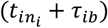 passed the first cut off time). In this case we call the resulting disease time the disease age. The setting is of the disease time is illustrated in Figure 5b & d. Using the above *T*_*b*_ means that the cut-off of the biomarker *Q*_1stb_ occurs at a disease age corresponding to the median {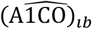 for *i* ∈ sample *b* with 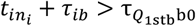}.

## 3. Results

### 3.1 Assessing the fit

Whether the results of the MMPP analysis are consistent with the assumption in equation (1) may be investigated by plotting the two quantities; the anchor times and mean trajectory curves from the MMPP analysis. Such a plot is shown in the last panel of Figure 1e, from which we can see the longitudinal data points are scattered closely around the trajectory curves, the extent of the scatter indicates how well the approximation holds. A measure of this scatter for the quantity *Q* is the mean square deviation from the trajectory per data point, 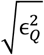 from equation (9), for simulated data set. The results of this successful application of the MMPP method are contrasted with an application on a simulated data set with two *independent* variables in section D in the supplementary material.

### 3.2 Results for disease trajectory curves

We constructed the multivariate disease curves for the simulated data using the five-step method and 1000 bootstrapped replicates to estimate the confidence limits, using the method from section 2.6 to fix the time shifts. The resulting disease trajectory curves are compared to the simulated data in Figure 6, which shows good agreement between the constructed curves and actual data.

**Figure 6:**
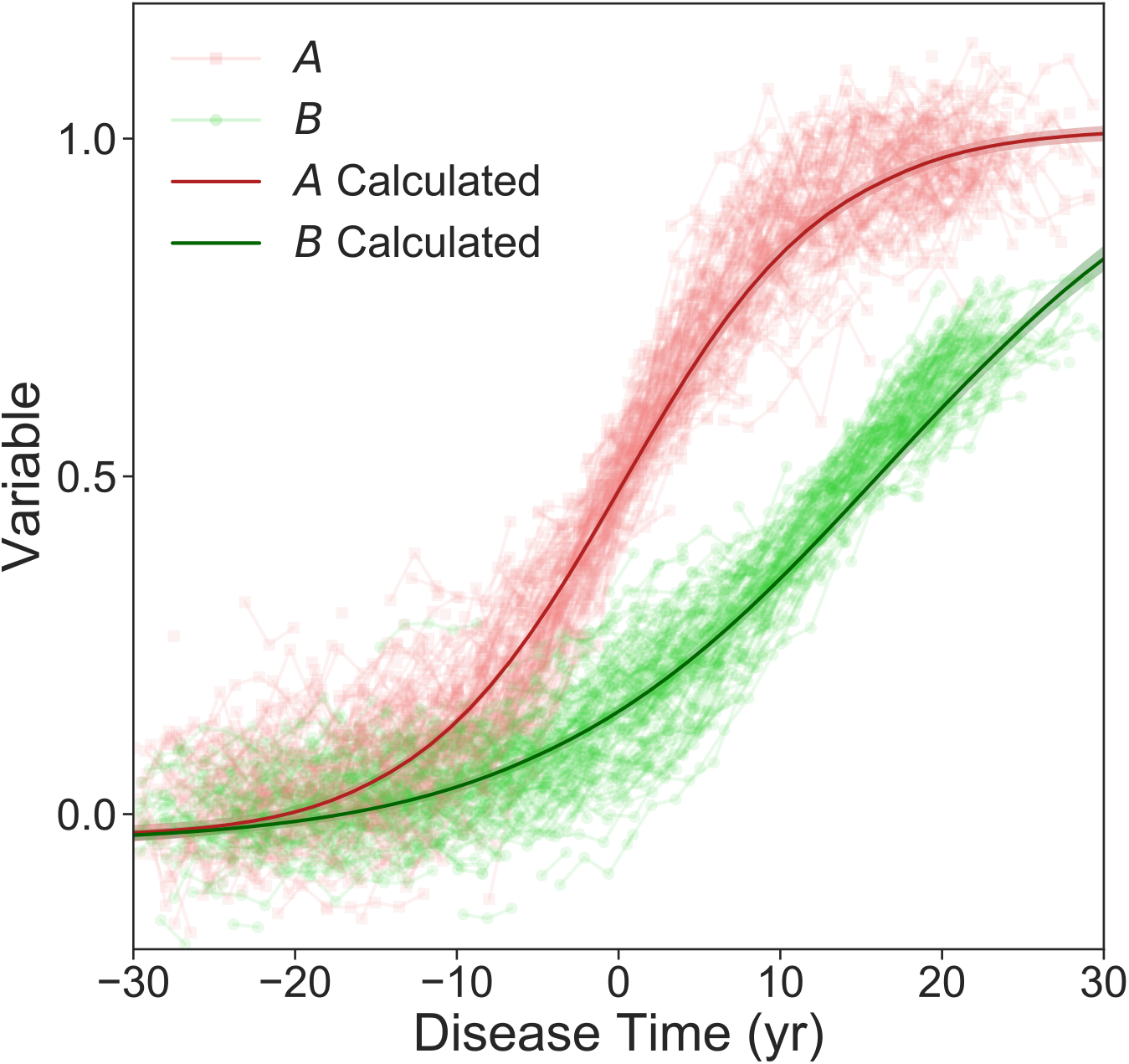
The MMPP estimates of the disease trajectory curves calculated with p=0.9 bounds shown compared to the simulated data as a function of its simulated disease time.

Quantities *A* and *B* are shown in red and green respectively.

We also calculated the delay times between variable *A* and variable *B* reaching values of 0.49 (the value halfway between the two asymptotic population values), both using the curves constructed with the MMPP technique and using the actual sampled curves 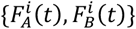. Using the MMPP technique we find the delay time 15.3 ± 0.4 yr (90% of bootstrapped curves give values in the range between the + and – values), in comparison to the actual sampled curves which have a delay time of 14 ± 2 yr. So, we can conclude the MMPP method has accurately estimated the delay time.

### 3.3 Individual anchor times

We also investigated how well the method determines the individual participant’s disease time. A plot showing calculated disease times for simulated individuals baseline collection and their simulated disease time for that point is shown in Figure 7a. The plot shows that for individuals with a baseline z-score of the *A* variable compared to the *t* → −∞ distribution (see section E of the supplementary material) *Z*_*A*_ ≳ 2; the simulated disease time is strongly predicted by the calculated disease time. We also performed a linear regression on the points with *Z*_*A*_ > 2 and found a straight line fit the data well with an *r*^2^ = 0.981. This shows that the disease times computed by MMPP give a reliable measure of disease stage for participants with *Z*_*A*_ > 2 in this example. One cannot expect the method to produce reliable disease times for participants whose *Z*_*A*_ ≲ 2 (which in this case implies *Z*_*B*_ ≲ 2), as in step 5 their data points will be indistinguishable from the pre-disease population and on the flat part of the disease trajectory and therefore hard to place accurately. We also produced a Bland-Altman plot shown in Figure 7b, for participants with *Z*_*A*_ > 2. From this plot we can conclude that the differences between MMPP estimates of the disease and the simulated disease time are essentially random for mean disease times ≳ −10 yr, or data with *Z*_*A*_ ≳ 2.6.

**Figure 7:**
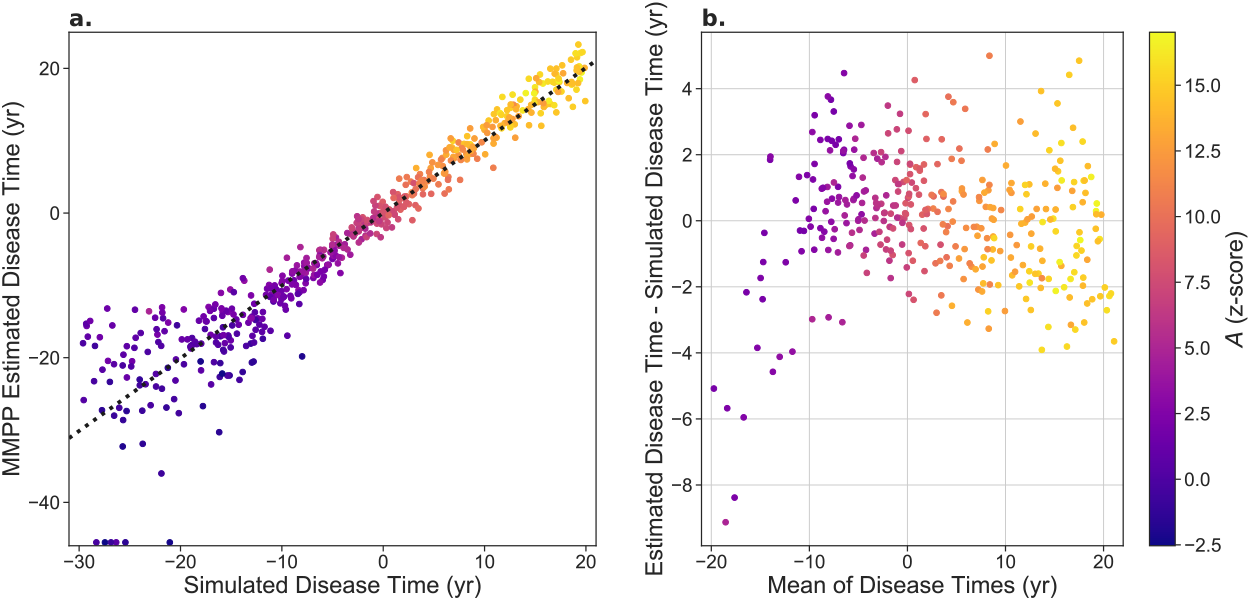
A comparison of the MMPP estimates for disease time with the simulated disease time. **a** The calculated disease time vs the simulated disease time for the simulated data set. The baseline data point for each simulated participant is shown. The dotted line shows a linear fit with of best fit to data with *Z*_*A*_ > 2 with *r*^2^ = 0.981. Only simulated participants with variable *A* increasing are shown in the plot. Note, there is a cluster of points with an estimated disease time of -55 yr because this is the lowest disease time we allowed in our minimiser. **b** A Bland-Altman plot comparing the disease times for data with *Z*_*A*_ > 2. In both plots points are coloured according to the *Z*_*A*_ value.

## 4. Discussion

Novel extensions to our phase-plane method [2] [7] for modelling longitudinal trajectories of disease progression is presented. The two main extensions are 1) true multivariate modelling of longitudinal trajectories of disease progression and 2) the ability to identify the placement of an individual on the multivariate trajectories of disease, in turn providing staging and prognostic information at the individual level.

We generate realistic simulated longitudinal data for individual trajectories, including point noise. This simulated data is then used to demonstrate that the MMPP method can be used to construct accurate estimates for the population level multivariate disease trajectories. Using the resulting disease trajectory curves, we produce accurate estimates of the delay times between different variables. At an individual level, we demonstrate that the MMPP method can accurately estimate the baseline disease time values for simulated participants when their earliest changing baseline measurement is significantly different (with a level p=0.98) from the pre/non-disease population. Using our bootstrapping methodology, we showed we were able to place confidence intervals for our disease curves and individual progression times.

The strengths of the updated method are as follows: the moving window method provides a more robust method for calculating sets of phase plane points from numerical longitudinal data. Further, the method also allows for consistent treatment of longitudinal data with missing data, different sampling rates between participants, and individuals with different numbers of data points. This is particularly important when applying the method to pooled datasets across different harmonized cohort studies.

The novel anchoring step, (step 5) allows multiple single variate disease curves to be aligned to one and other, even when different levels of noise are associated with different variables. Unlike other methods [5], the full MMPP when properly implemented does not require *a priori* selection of one variable to anchor the others, both when constructing the curves and placing individuals on the curves. Multivariate data from participants who do not have enough visits for an accurate estimate for the rate of change to be estimated can still be used to contribute to the anchoring step. Also, by changing the fit function in step 2 one can construct disease trajectory curves for variables that follow trajectories which could be very different than the sigmoid like trajectories in our simulated data.

There are several limitations that need to be kept in mind when applying the MMPP; we believe some could be mitigated with further research. Firstly, it remains to be seen how well the method predicts future individual outcomes for longitudinal study participants, however the method is not designed for prediction. Secondly, the bootstrap method does not give estimates for how far individuals’ data may deviate from the population level curves, and different methods will need to be used to quantify this. We have not investigated the effectiveness of different bootstrapping methodologies applied to the MMPP, so it is possible there is a better method than the one presented here. Finally, the MMPP method is still susceptible to the selection bias inherent in the longitudinal studies on which it is applied.

It should be noted that the main assumptions about AD used in constructing the simulated data set are: (a) that the variables have well defined progression curves over the course of the disease (i.e. that satisfy the approximation in equation (1)) and (b) that the age of onset of biomarker change varies from person to person. Hence, this method may find some application studying other diseases for which the assumptions hold such as Parkinson’s disease, multiple sclerosis or certain cancers. While the MMPP method is not directly applicable to variables that are static or without well-defined progression curves, it may be interesting to study how these variables effect the disease times of study participants.

In conclusion, we develop here a method to construct multivariate disease trajectories from longitudinal data sets for AD. The method also accurately places study participants on the disease progression curves to provide individualised staging. The method presented here has many possible applications considering the plethora of new biomarkers that have been discovered. In particular, it is vital to understanding the timing of changes in different biomarkers measuring Aβ and tau (eg. imaging, blood-based biomarkers and CSF) and their relation to cognitive performance and disease progression.

## Data Availability

All data produced in the present study are available upon reasonable request to the authors

