## Supplementary material for "A multi-modal phase plane method for constructing multivariate disease trajectories": Supp

#### A: evaluating $\dot{\mu}_Q^*(\tau - \tau_{Q0})$ .

The differential equation

$$\dot{\mu}_Q^* = f_Q\left(\mu_Q^*\right), \qquad (A1)$$

Can be reduced to quadrature by separation of variables,

$$\tau - \tau_{Q0} \coloneqq \Delta\tau\left(\mu_Q^*(\tau)\right) = \int_{\mu_Q^*(\tau_{Q0})}^{\mu_Q^*(\tau)} \frac{\mathrm{d}q}{f_Q\,(q)}. \qquad (A2)$$

$\mu_Q^*\left(\tau_{Q0}\right)$ is a value that we chose which is in the biologically relevant (in applications we would choose $\mu_Q^*\left(\tau_{Q0}\right)$ to be the threshold value for abnormality of the variable Q or some other significant milestone). To produce values of $\dot{\mu}_Q^*(\tau - \tau_{Q0})$ rapidly we follow the procedure: (i) identify the region values of Q for which we need the curve, this will either a region encompassing all the longitudinal data, or if $f_Q\,(q)$ has roots (which will be asymptotic values of the curve $\mu_Q^*(\tau - \tau_{Q0})$) in this range the region containing our reference value starting a small distance $\delta_Q$ from the root (for example if $f_Q\,(Q) = Q(2 - Q)$, which has roots at Q=0,2 and we had data between -0.2 and 1.5 a reference value of $\mu_Q^*\left(\tau_{Q0}\right) = 0.5$ and $\delta_Q = 10^{-5}$ the region could be chosen $10^{-5}$ to 1.6). (ii) divide the region into a grid of $\mu_Q^*(\tau)$ values with a suitable spacing (in the example in the text we divided the region into a grid of n=200 equally spaced points) and compute $\Delta\tau\left(\mu_Q^*(\tau)\right)$ on this region using Simpson's rule for approximation of the integral (A2). (iii) then we would have a grid of $\{\mu_Q^*(\tau_{(Qk)})\}_{k=1}^{\mathrm{n}}$ values with associated $\{\tau_{(Qk)} - \tau_{Q0}\}_{k=1}^{\mathrm{n}}$ values and values for $\mu_Q^*(\tau)$ for arbitrary $\tau$ inside the range $(\tau_{(Q1)}, \tau_{(Q\mathrm{n})})$ we compute using a piecewise cubic Hermite interpolating polynomial of this grid of values, (this choice of interpolation ensures the approximation for $\mu_Q^*(\tau)$ is monotonic and sufficiently smooth for the BFGS minimisation algorithm). If values outside this range are ever needed, they can be computed by the more computationally intensive method of solving the differential equation with a stiff solver starting at the appropriate end of the grid. To ensure that the multivariate trajectories for the full set of curves are computed on the same range of disease times, we add the values $T_{min} = \min_{Q\in\mathcal{Q}}(\{\tau_{(Q1)}\})$, $T_{max} = \max_{Q\in\mathcal{Q}}(\{\tau_{(Q\mathrm{n})}\})$, and associated $\mu_Q^*(\tau_{(Qk)})$ values to the grid's for biomarkers which do not have values at these disease times by integrating the differential equations.

### B: Initial guess for solver.

In order to provide an initial guess to the solver described in the text we need initial values for $|\mathcal{Q}| - 1$, $\tau_{Q0}$'s and $N$ $\tau_i$'s, (one of the $\tau_{Q0}$ is set $\tau_{Q0} = 0$). We call these guesses $\{\tau_{Q0}{}^g\}$ and $\{\tau_i{}^g\}$. In practise if one of the variables is known a priori to be less noisy then (unless there is another variable which changes much earlier), this variable should have $\tau_{Q0} = 0$ and if not the minimisation procedure should be run once for each possible choice and the minimum final negative-log likelihood should be chosen. Once we have made this choice for $\tau_{Q0} = 0$, call the fixed variable P, then for each participant i chose the value of for the initial guess of $\tau_i{}^g$ we take the value such that aligns participant i's mean P with the curve,

$$\mu_P(\tau_i{}^g) = \overline{P_i}.$$

Where $\overline{P_i} = \sum_{m=1}^{n_i} P_{im}/n_i$ is the participants mean value for the biomarker P. For participants whose $\overline{P_i}$ is outside of the range of $\mu_P(\tau)$ we either use a regularised version of $\mu_P(\tau)$ (see next section) or set $\tau_i{}^g = \pm T_{\text{Big}}$, where $T_{\text{Big}}$ is a large value (say 60 y) and the +/- is chosen depending on whether whose $\overline{P_i}$ is above/below the range of $\mu_P(\tau)$. Then if any of the $|\tau_i{}^g| > T_{\text{Big}}$ then set those to $\tau_i{}^g = 0$ to remove any extreme values (the minimisation algorithm should be robust to a number of participants starting a disease time that does not align with the P curve).

Once we have a guess for every individual anchoring we can compute initial guesses for $\tau_{Q0}, (Q \neq P)$ by minimising the squared sum of residual in the variable of Q, that is chosing $\tau_{Q0}{}^g$ which minimises

$$\mathcal{E}_Q^2\left(\{\tau_i{}^g\}_{i\leq N}, \tau_{Q0}{}^g\right),$$

which we compute again using the SciPy implementation of the (BFGS) quasi-Newton algorithm, with an intitial guess for $\tau_{Q0}{}^g$ of 0.

### C: Regularising the minimisation problem.

In cases like the simulated data set in the main text and real amyloid β accumulation problems the minimisation problem is non-identifiable, because participants who have biomarker values indistinguishable from the asymptotic values can, with negligible change in likelihood be anchored at any point sufficiently far along the asymptote. Biologically we cannot expect to be able to place these participants accurately with only the biomarker information available. We make the problem identifiable by regularising it, we examined to two approaches of regularising the problem: (i) regularising the curves and (ii) regularising the pseudo likelihood.

### C.1 Regularising the curves.

To regularise the curves, we make the curves go well outside of the range of data at a distance down any asymptotes so that the negative log-likelihood cost for placing anchoring participants to far along the asymptotes. In practice we do this by adding points to our interpolation grid an arbitrary $\Delta T = 1y$ away from any asymptotes with values an arbitrary large number $\Lambda$ (at least be comparable the range of our data for the example we use $\Lambda = 10$) added or subtracted from to the markers asymptotic value. This modification of the curve illustrated for a fictional example in supplementary figure C1.

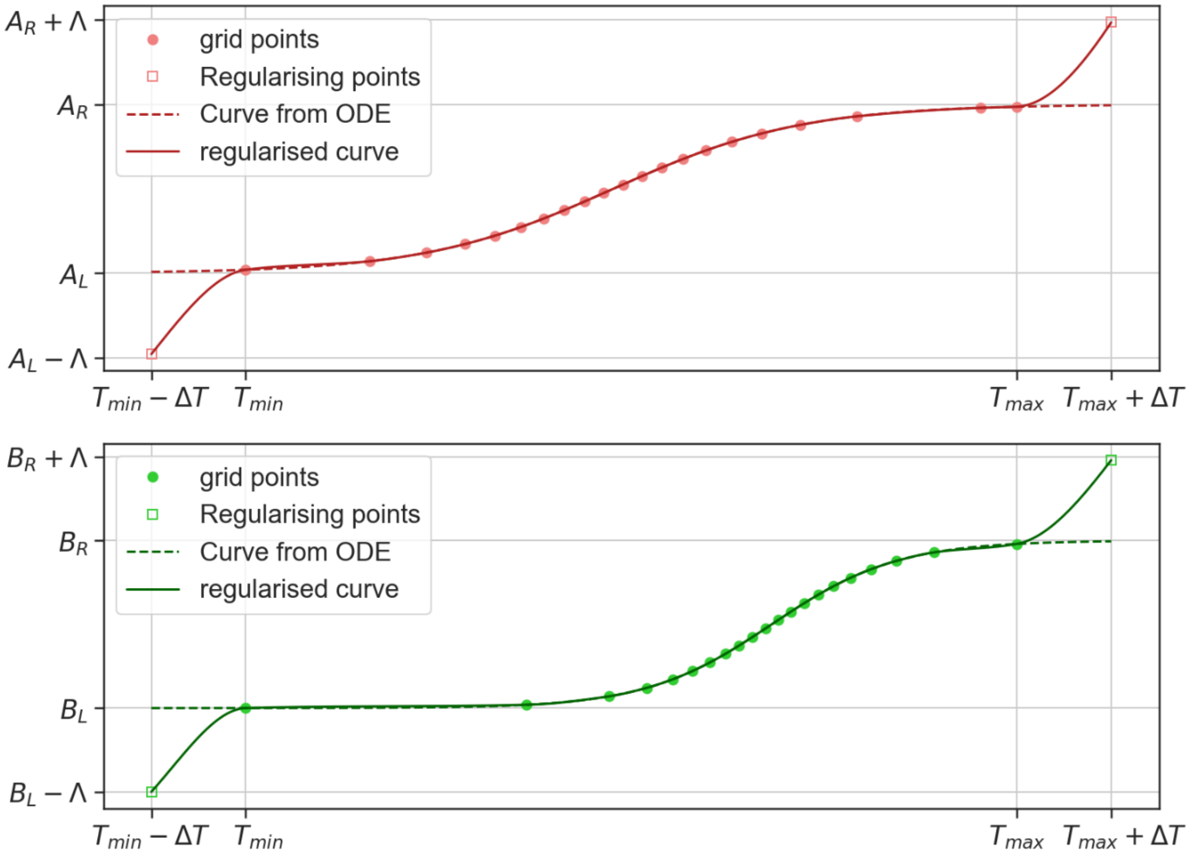


*Figure C1 producing regularised curves, we add two regularizing points (square markers) as described in the text, to our interpolation grid (circular markers) to produce regularised curves (solid lines) by interpolation, compaired ot the unregularized curves obtained by solving the ordinary differential equation (ODE) A1 shown with a dashed line. The curves are shown for two quantities A &B having left/right asymptotes at $A_L/A_R$ and $B_L/B_R$ respectively.*

### C.2 Regularising likelihood.

In this approach we add a term $-\ell_{\mathrm{Reg}}$ to the negative log likelihood (equation $(6)$ in the main text) which penalises extreme values for the anchoring times, this cuts off the large regions of constant likelihood at asymptotes. Any increasing function which is approximately zero in the range of interest and grows smoothly and rapidly outside this range could be used for $-\ell_{\mathrm{Reg}}$ we use the following

$$-\ell_{\mathrm{Reg}} = \frac{1}{N + |\mathcal{Q}| - 1} \sum_{\tau_a \in \mathcal{T}} \left(\frac{\tau_a}{T_{\mathrm{Big}}}\right)^2 \tanh^4\left(\frac{\tau_a}{T_{\mathrm{Big}}}\right),$$

in this expression the sum is taken over the set $\mathcal{T} = \{\tau_i\}_{i=1}^{N} \cup \{\tau_{Q0}\}_{Q \in \mathcal{Q}, Q \neq P}$ of all $N + |\mathcal{Q}| - 1$ anchor times which are fit and $T_{\mathrm{Big}}$ is a large time period (we use $T_{\mathrm{Big}} = 100y$). The function $\left(\frac{\tau}{T_{\mathrm{Big}}}\right)^2 \tanh^4\left(\frac{\tau}{T_{\mathrm{Big}}}\right)$ is plotted in figure C2.

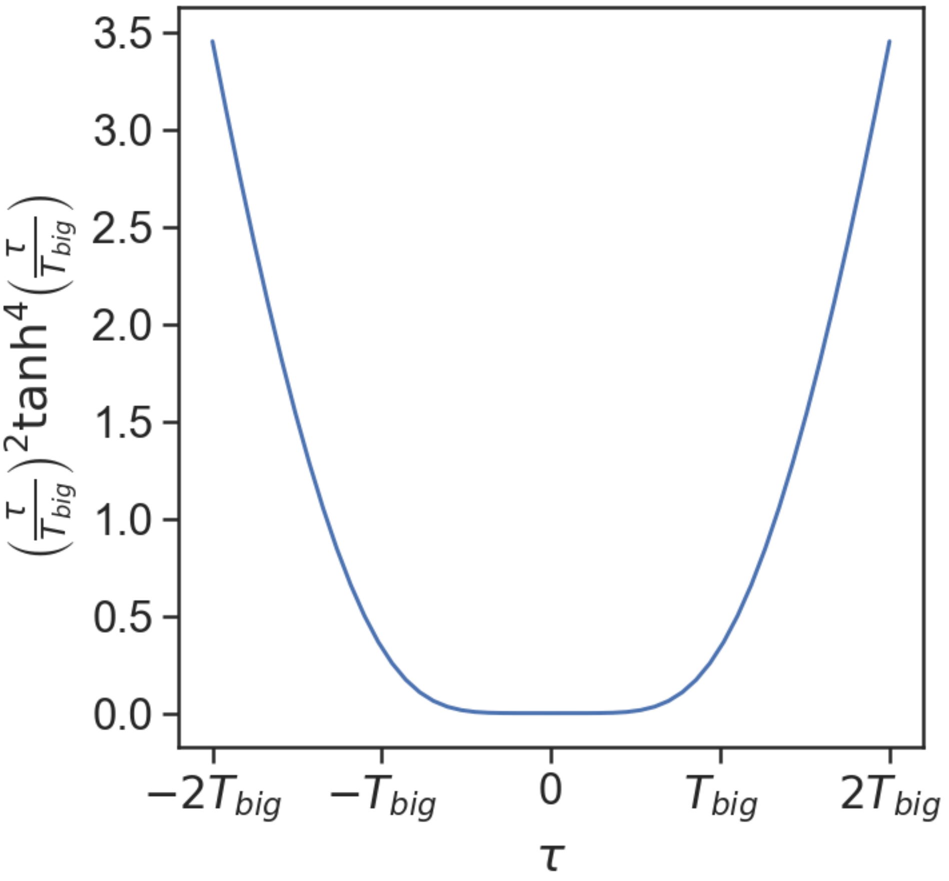


*Figure C2 a plot of the function* $\left(\frac{\tau}{T_{Big}}\right)^2 tanh^4\left(\frac{\tau}{T_{Big}}\right)$

### D: variables with uncorrelated disease progression

In order to better understand when the method fails, we created a simulated data set where the disease time shift between the two variables was more widely varying. In this *independent progression* simulation, data for the simulated variables *A* and *B* was generated exactly as described in section 2.2 of the main text except the time lag between the two variables was drawn from a wider distribution

$$\Delta b_B^i \sim \mathcal{N}(0,100^2) \qquad \text{(independant progression)},$$

when compared to the simulation in the text with

$$\Delta b_B^i \sim \mathcal{N}(14,0.1^2) \qquad \text{(section 2.2)}.$$

The simulated independent progression data set is shown in figure D1. When the MMPP method is applied to the independent progression data set we see that the constructed disease trajectories do not well approximate the data in the independent progression data set like they do to the simulated data from section 2.2. The mean squared deviation for this data set are $\sqrt{\epsilon_A^2}$ =0.12 and $\sqrt{\epsilon_B^2}$ =0.14.

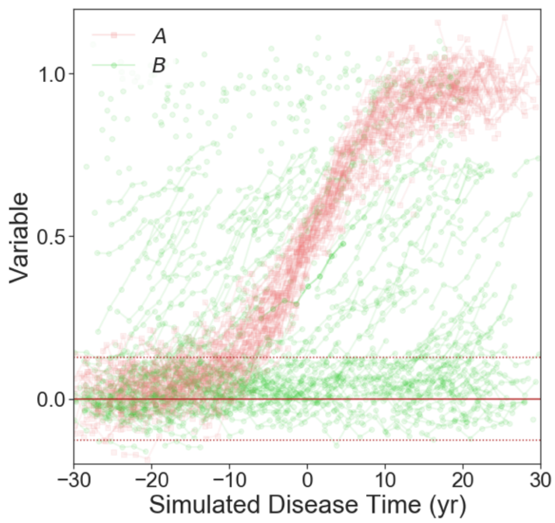


Figure D1: The full simulated longitudinal independent progression data set for the variables *A* and *B* in plotted as a function of the simulated disease time variable, the simulated pre-disease mean of the *A* variable is shown with a solid line, and the two standard deviations spread of the pre-disease population are marked with dotted lines.

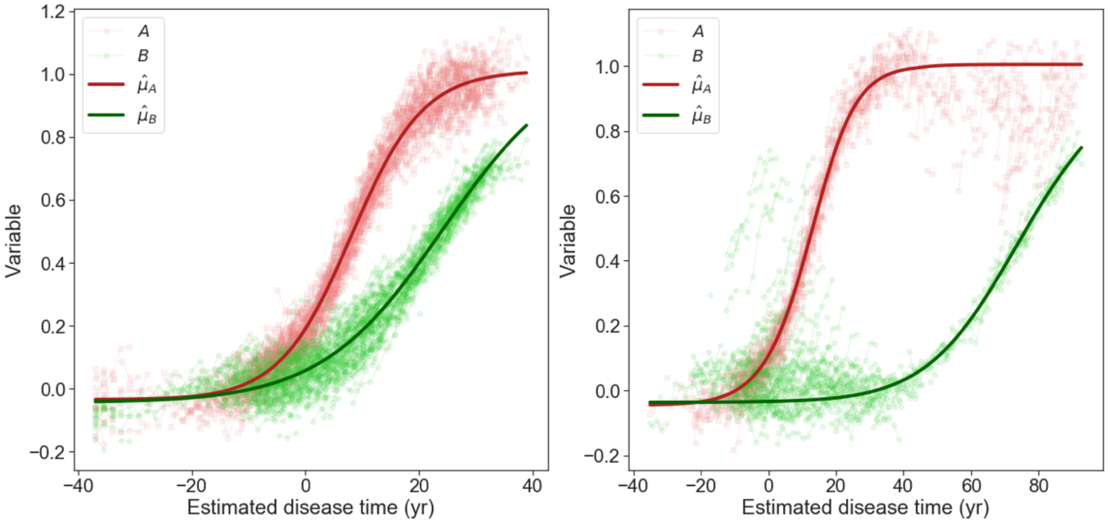


Figure D2: Disease time plots for both the simulated data set discussed in the main text (left) and the independent progression data set (right). Points show the anchored longitudinal data and the lines show the MMPP estimated trajectories.

### E: The distribution of the simulated variables in a pre-disease population

The distribution for the simulated variables *A* and *B* in a pre-disease population can be obtained by taking the limit: $t \to -\infty$. This means for our example dataset the pre-disease distribution of *A* and *B* is

$$\begin{aligned} A &\sim \mathcal{N}(0, 0.0707^2) \\ B &\sim \mathcal{N}(0, 0.0510^2). \end{aligned}$$

This allows us to compute z-scores $Z_A$ and $Z_B$ of the variables with respect the pre-disease distribution.
